# A snap shot of space and time dynamics of COVID-19 risk in Malawi. An application of spatial temporal model

**DOI:** 10.1101/2020.09.12.20192914

**Authors:** Alfred Ngwira, Felix Kumwenda, Eddons Munthali, Duncan Nkolokosa

## Abstract

**Background:** COVID-19 has been the greatest challenge the world has faced since the second world war. The aim of this study was to investigate the distribution of COVID-19 in both space and time in Malawi.

**Methods:** The study used publicly available data of COVID-19 cases for the period from 24^th^ June to 20^th^ August, 2020. Semiparametric spatial temporal models were fitted to the number of weekly confirmed cases as an outcome data, with time and location as independent variables.

**Results:** The study found significant main effect of location and time with the two interacting. The spatial distribution of COVID-19 showed major cities being at greater risk than rural areas. Over time the COVID-19 risk was increasing then decreasing in most districts with the rural districts being consistently at lower risk.

**Conclusion:** Future or present strategies to avert the spread of COVID-19 should target major cities by limiting international exposure. In addition, the focus should be on time points that had shown high risk.

## Introduction

COVID-19 is a corona virus disease (COVID which was first reported in Wuhan, China in 2019. It is characterized by severe acute respiratory syndrome (SARS) hence also known as SARS-COV − (WHO, 2020). Since its onset, COVID-19 has been one of the greatest disease pandemics of all times. From its discovery in December in China, 19 718 030 people world over have been confirmed of the disease and over 722 285 people have died as of 9 August 2020 (WHO, 2020). The space trend worldwide has shown the Americas (10 590 929), Europe (3 3 582 911) and South East Asia (2 632 773) being the hardest hit as of 10^th^ August, 2020. Africa had a slow progression of the disease at the beginning of the disease, earlier 2020, but the continent had rising cases in the middle of the year, 2020, with 895 696 confirmed cases and 16 713 deaths as of 10^th^ August, 2020. Malawi at this time of the year had recorded 5193 confirmed cases and 163 deaths (UNICEF Malawi, 2020b). The first three cases of COVID-19 were recorded on 2^nd^ April, 2020 (UNICEF Malawi, 2020a).

Understanding disease space and time dynamics is important for the epidemiologists as with space distribution, the hot spot areas are marked for intervention. In addition, possible drivers of the epidemic in those hot spots are suggested for further scientific investigation. Regarding temporal distribution, times with high disease risk are also identified which gives crew to possible causes including, in particular, seasonal changes. A number of studies on spatial temporal distribution of COVID-19 have been conducted (Chen et al, 2020; Yang et al, 2020; Gayawan et al, 2020; Martellucci et al, 2020; Adekunle et al, 2020, Arashi et al, 2020; Kumar et al, 2020; Briz-Redón and Serrano-Aroca, 2020; Diop et al, 2020; Jia et al, 2020; Xie at al, 2020; Likassa, 2020; Sarfo and Karuppannan, 2020; Daw et al, 2020; Ye and Hu, 2020). The majority of these though have used the geographical information system (GIS) technology as compared to statistical modelling using spatial temporal models. A few studies that have used the statistical approach to spatial temporal analysis to my knowledge are Gayawan et al (2020) who used the Possion hurdle model to take into account excess zero counts of COVID-19 cases, Briz-Redon and Serrano Aroca (2020) who used the separable random effects model with structured and unstructured area and time effects, and Chen et al (2020) who used the inseparable spatial temporal model. In addition, in Africa, spatial temporal analysis of COVID-19 cases has been limited (Gawayan et al, 2020; Arashi et al, 2020; Adeneknle et al, 2020) as of 9^th^ August. In Malawi, at the time of this study, no study on spatial temporal distribution of COVID-19 cases had been spotted. Only one study that focused on prediction of COVID-19 cases using mathematical models was seen (Kuunika, 2020). The aim of this study was to determine the spatial temporal trends of COVID-19 confirmed cases in Malawi while using the spatial temporal statistical models. The objectives of the study were:

- To establish the estimated or predicted risk trend by geographical location
- To estimate the temporal risk trend of COVID-19 by geographical location.

The article has been organized as follows. First, study methods are described in terms of data collection and statistical analysis. Thereafter, results and discussion of results are presented. Finally conclusions and implications regarding the findings of the study are made.

## Materials and Methods

### Data

The study used the publicly available districts’ COVID-19 confirmed daily cases data for Malawi which was extracted from the Malawi data portal website (https://malawi.opendataforafrica.org) after registering with the portal. The total population, population density, and percentage of people with running water data for each district were also extracted from the same data portal. The population size for each district was used as the expected number of people to be infected in each district. Population density and percentage of people with running water in each district were taken as covariates. Though the cases started to be recorded on 2^nd^ April, 2020, the extracted data for COVID-19 cases used for spatial temporal modelling in this study, only covered the period from 24^th^ June to 20^th^ August. This was the case, considering that COVID-19 daily cases for the districts were only available from 24^th^ June on the portal. The study period was divided into six weeks as follows: week 1 (24/06/20 – 02/07/20), week 2 (02/07/20 – 09/07/20), week 3 (09/07/20 – 16/07/20), week 4 (16/07/20 – 23/07/20), week 5 (23/07/20 – 11/08/20) and week 6 (11/08/20 – 20/08/20). The daily cases were then aggregated at week level to make week the unit of analysis for time. The Malawi data portal is a web based data storage system that displays data on main indicators of the country. No any other permission was required to publish the results of the analysis.

### Statistical analysis

Descriptive analysis involved the time series plot of the cumulative confirmed cases and those who had died of COVID-19 for the whole country from the beginning of the epidemic to the time this study was conducted, that is, 20^th^ August, 2020. It also involved bivariate correlation of daily confirmed cases of COVID-19 and their potential covariates, that is, population size, population density and percentage of people with running water in each district. The potential covariates would be selected for further multiple variable modelling if their p-values were less than 0.20. After the descriptive analysis, the multiple variable spatial temporal models were fitted in R using the Bayesian approach while using the integrated nested laplace approximations (INLA). INLA estimation of Bayesian hierarchical models is proved to be faster than the use of Markov Chain Monte Carlo (MCMC) methods. A spatial temporal model assuming linear trend of cases over time as proposed by (Bernardinelli et al, 1995) was first fitted. That is, let y_*ij*_ be the observed disease cases in region *i* and at time *t*, then the observed disease cases have the Poisson distribution with the data model defined as *y_it_* ∼ Poisson (*e_it_r_it_*), where*e_it_r_it_* is the mean of disease cases, *e_it_* and *r_it_* are the expected disease cases and relative risk respectively. The model of the relative risk is then defined as log(*r_it_*)=η_it_ where η_it_ is the predictor specified as

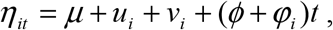

The model with predictor (1) will be denoted by A. In (1),*μ* is the overall disease relative risk, *u _i_* is the area level unstructured random effect and *v_i_* is the area level structured random effect, and *ϕ* is overall time trend and *φ_i_* is the area specific time trend. The unstructured area level effects were modelled by the independent normal distribution with zero mean, that is, 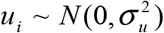, and the structured random effects were assigned the intrinsic conditional auto-regressive (ICAR) according to Besag (1991), that is, 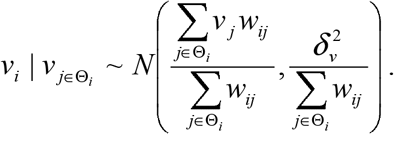.The weakness of model A is the linearity assumption on the effect of time on the relative risk of the disease. To take a more flexible approach on the effect of time, nonlinear spatial temporal models were also explored. The predictor,*η_it_* for the nonlinear spatial temporal model for the time effect is specified as follows:

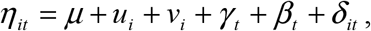

and let it be denoted by B. The *u_i_* and *v_i_* in the model are the area level unstructured and structured effects respectively as defined in (1), and *γ_t_* and *β_t_* are the unstructured and structured temporal effects. The unstructured time effects was modelled by the independent normal distribution with zero mean, that is, 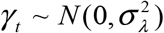 and the structured temporal effects were assigned the first order random walk prior distribution defined as:
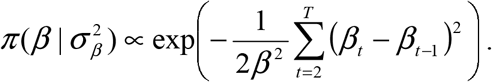. A second order random walk was also explored in case the data would show a more pronounced linear trend. The second order random walk is defined as: 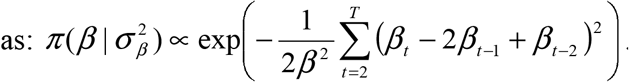. The last term in (2),*it_δ_*, represents the interaction between area and time. Four forms of interaction between space and time are possible according to Knorr-Held (2000). The first form of interaction assumes interaction between the unstructured region effect (*u_i_*) and the unstructured temporal effect (*γ_t_*) (denote it by model B 1), and in this case the interaction effect is assigned the independent normal distribution, that is, 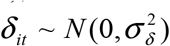. The second type of interaction, is the interaction of structured area effect (*v_i_*) and the unstructured temporal effect (*γ_t_*)(denote it by model B 2). This form of interaction assumes conditional intrinsic (CAR) distribution for the areas for each time independently from all the other times. The third is the interaction between unstructured area effect (*u_i_*) and the structured temporal effect (*β _t_*)(denote this by model B 3). The prior distribution for each area is assumed to be a second order random walk across time. The last possible space time interaction is that between area structured effect (*v_i_*) and the structured time effect (*β_j_*)(call it model B 4). In this case, the second order random walk prior that depends on neighboring areas was assigned for each area. The prior for the variance parameter for the independent normal distribution (i.i.d) and that for the spatial effect (Besag) was the gamma (1, 0.001). The prior for the random walk variance was the gamma (1, 0.0005). The intercept was assigned the default normal, N (0, 0). The model choice was by the deviance information criteria (DIC) as proposed by Spiegelhalter et al (2002), where a smaller DIC means a better model in terms of fit and complexity. It is the sum of the measure of model fit, called the deviance denoted by 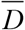 and the effective number of parameters denoted by 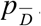. The selected model was then used to estimate the relative risk,*r_it_*.

## Results

Figure 1 shows the graph of cumulative confirmed and dead cases of COVID-19 from the time the first case was reported to 20^th^ August, 2020. There were 5282 confirmed cases and 165 deceased cases as of 20^th^ August, 2020. Generally, there were low total cases of those who died of COVID-19 as compared to those who were confirmed.

**Figure 1:**
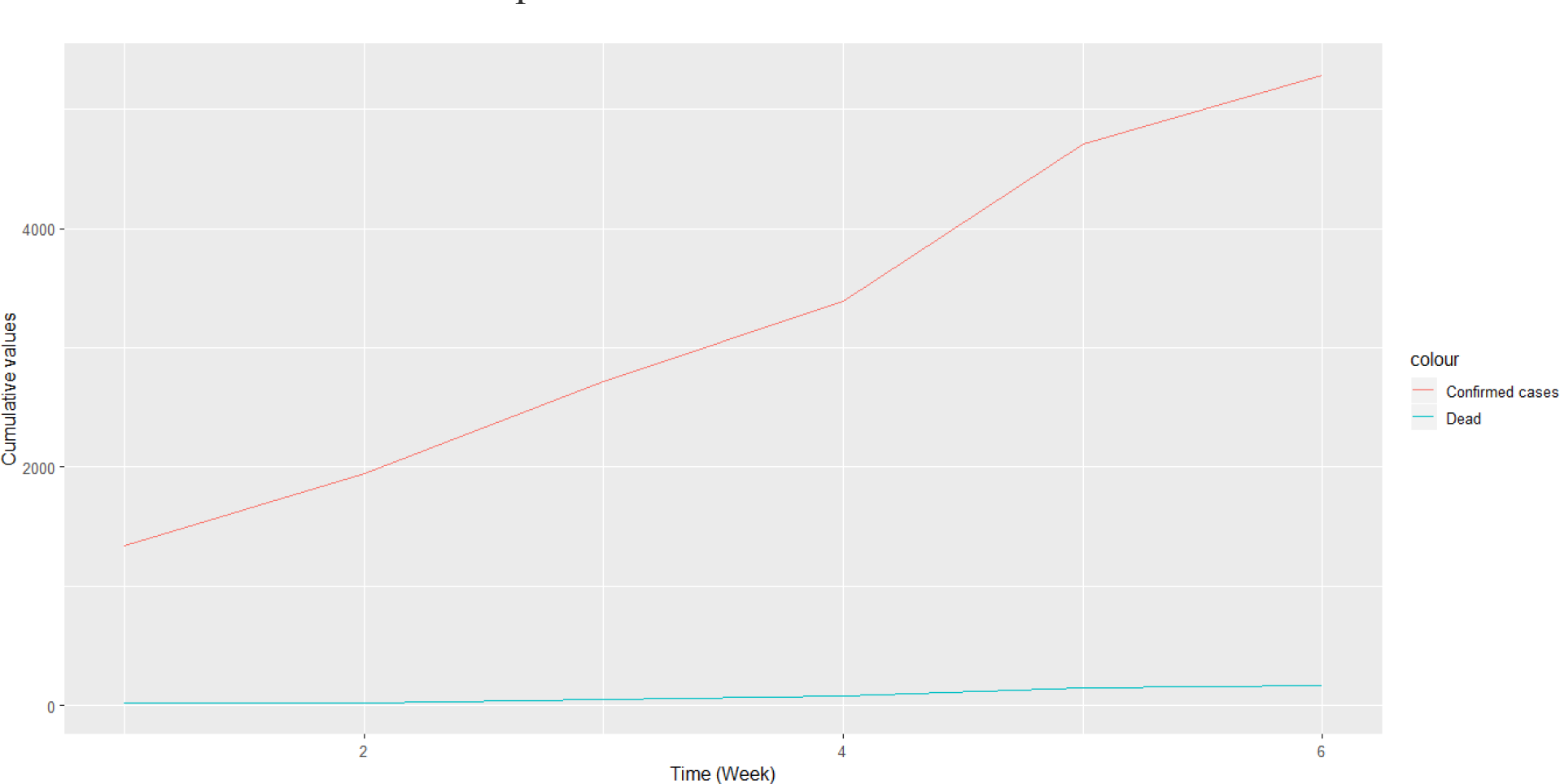
Cumulative cases of confirmed and those who had died of COVID-19 in Malawi from 24/06/20 to 20/08/20.

The pairwise Spearman correlation between the confirmed cases and the population density (*ρ*=−0.065, p-value = 0.380), revealed lack of correlation. There was also no significant correlation between confirmed cases and percentage of people with running water in each district (*ρ*=0.000, p-value = 0.998). Since the p-values of the correlation coefficients were more than 0.20, the significance level set to select potential covariates, the two covariates, population density and proportion of those with running water were dropped when fitting the spatial temporal models of the weekly confirmed cases of COVID-19.

Table 1 shows the DIC of the fitted spatial temporal models defined in the methods section. Model B 11 and B 12, had smaller DIC than the rest of the models, and the DIC difference between the two models was not significant as it was not greater than 3 (Spiegelhalter et al, 2002). The results of the model with RW2, that is, Model B 11 are therefore presented and discussed. Table 2 presents the variance parameters of the random effects. All model terms including the interaction effect of location and time, were significant predictors of the risk of contracting COVID-19 as the estimated variances were significantly greater than zero, since the confidence intervals excluded zero. Area level spatial effects and the effects of time modelled by the second order random walk prior (RW2), were highly significant as evidenced from their bigger variances.

**Table 1:**
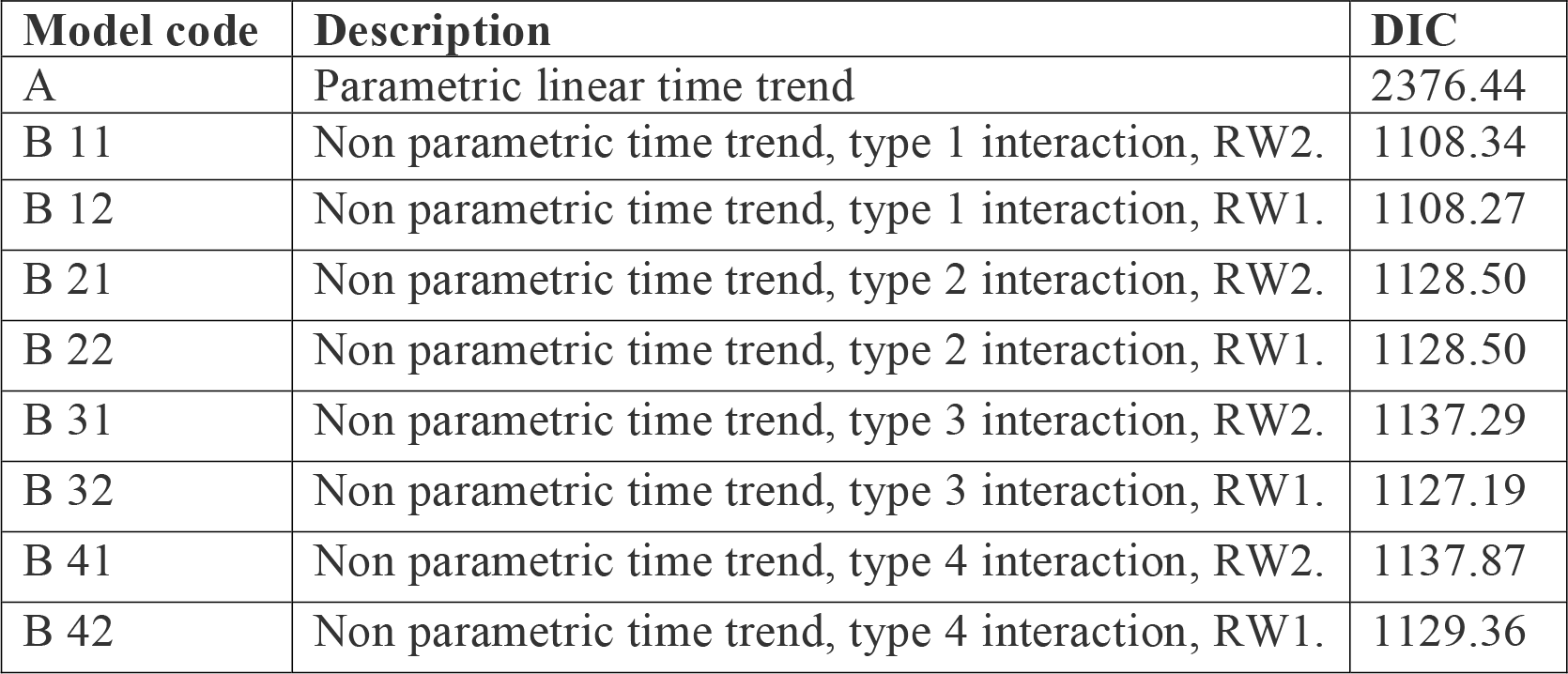
DIC of all fitted models

**Table 2:** Variance parameters of model B 11

| Variable | $\sigma^2$ (95 % Confidence Interval) |
| --- | --- |
| Area (i.i.d) | 0.49 (0.27, 0.77) |
| Area (spatial) | 1818.32 (128.30, 6657.27) |
| Time (i.i.d) | 13.62 (2.57, 43.43) |
| Time (RW2) | 20089.19 (1189.52, 70846.15) |
| Area x Time (i.i.d) | 3.29 (2.33, 4.58) |

The spatial temporal distribution of overall fitted risk (Figure 2, Figure 3), shows that by space, Mzimba, Mzuzu and Nkhata Bay in the north; Lilongwe, Lilongwe City and Mchinji in the center; Blantyre, Mwanza, Zomba and Mangochi in the south; were at increased risk to being confirmed of COVID-19. Overtime, from Week 1 to Week 6, the risk in Mzimba and

Mzuzu, had been decreasing and the risk in Blantyre was consistently high. Most of the districts in the rural areas were consistently at low risk of contracting COVID-19 over time.

**Figure 2:**
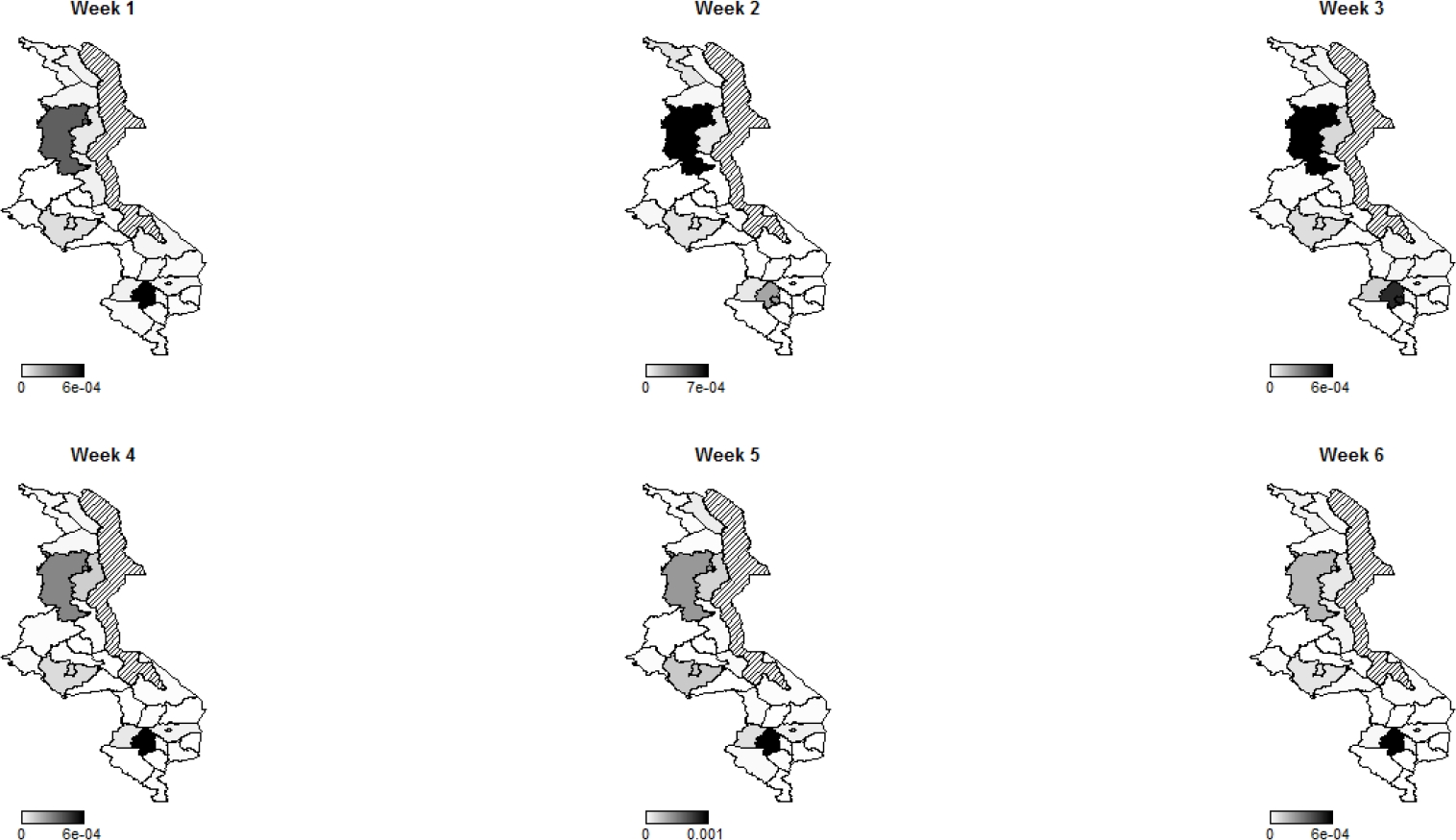
Evolution of overall predicted risk by district over time. Dark (high risk), and white (low risk).

**Figure 3:**
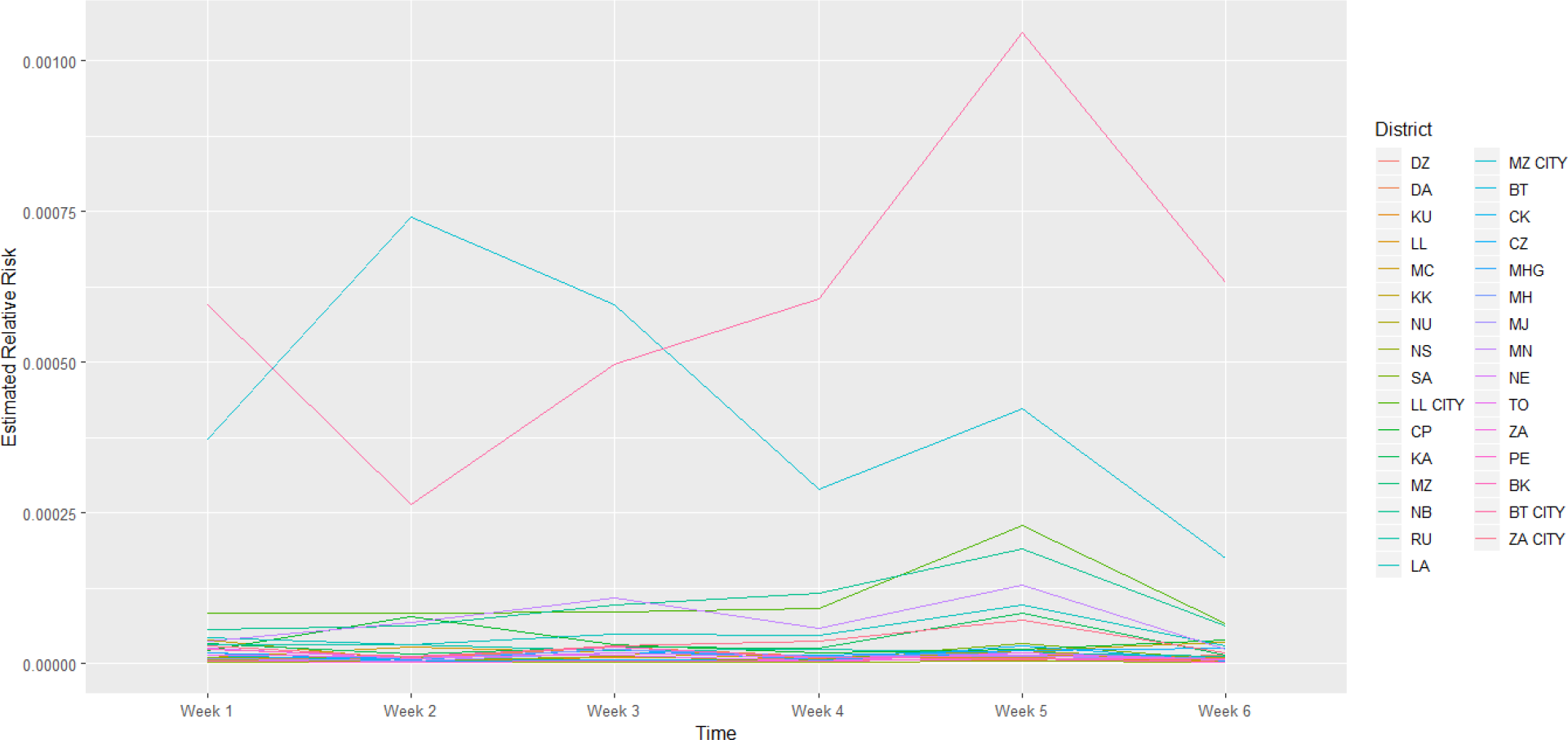
Evolution of relative risk by district over time. DZ: Dedza; DA: Dowa; KU: Kasungu; LL: Lilongwe; MC: Mchinji; KK: Nkhota-Kota; NU: Ntcheu; NS: Ntchisi; SA: Salima; CP: Chitipa; KA: Karonga; MZ: Mzimba/Mzuzu; CK: Chikwawa; CZ: Chiradzulu; MHG: Machinga; MH: Mangochi; MJ: Mulanje; MN: Mwanza; NE: Nsanje; TO: Thyolo; ZA: Zomba; PE: Phalombe; BK: Balaka.

Figure 4 presents the spatial risk of contracting COVID-19. Spatial risk represents the resdual risk due to unobserved or unmeasured factors of COVID-19. In general, the spatial risk looks randomly distributed in week 1, 3, 4 and 5 and non-random in week 2 and 6. Precisely though,in the first week, the risk was high in the south than in the center and north. In the second week, the spatial risk was high in the north and a little bit in the center close to the western border, than in the south. In the third week, the spatial risk was high in Dowa (center) and south of the lake, that is, Machinga and Mangochi. In the fourth week, the spatial risk was high in areas surrounding major cities in all the three regions. In the fifth week, the spatial risk shifted to the south and to one district in the northern region. Most areas in the last week, had reduced spatial risk to COVID-19, with exception to two central districts.

**Figure 4:**
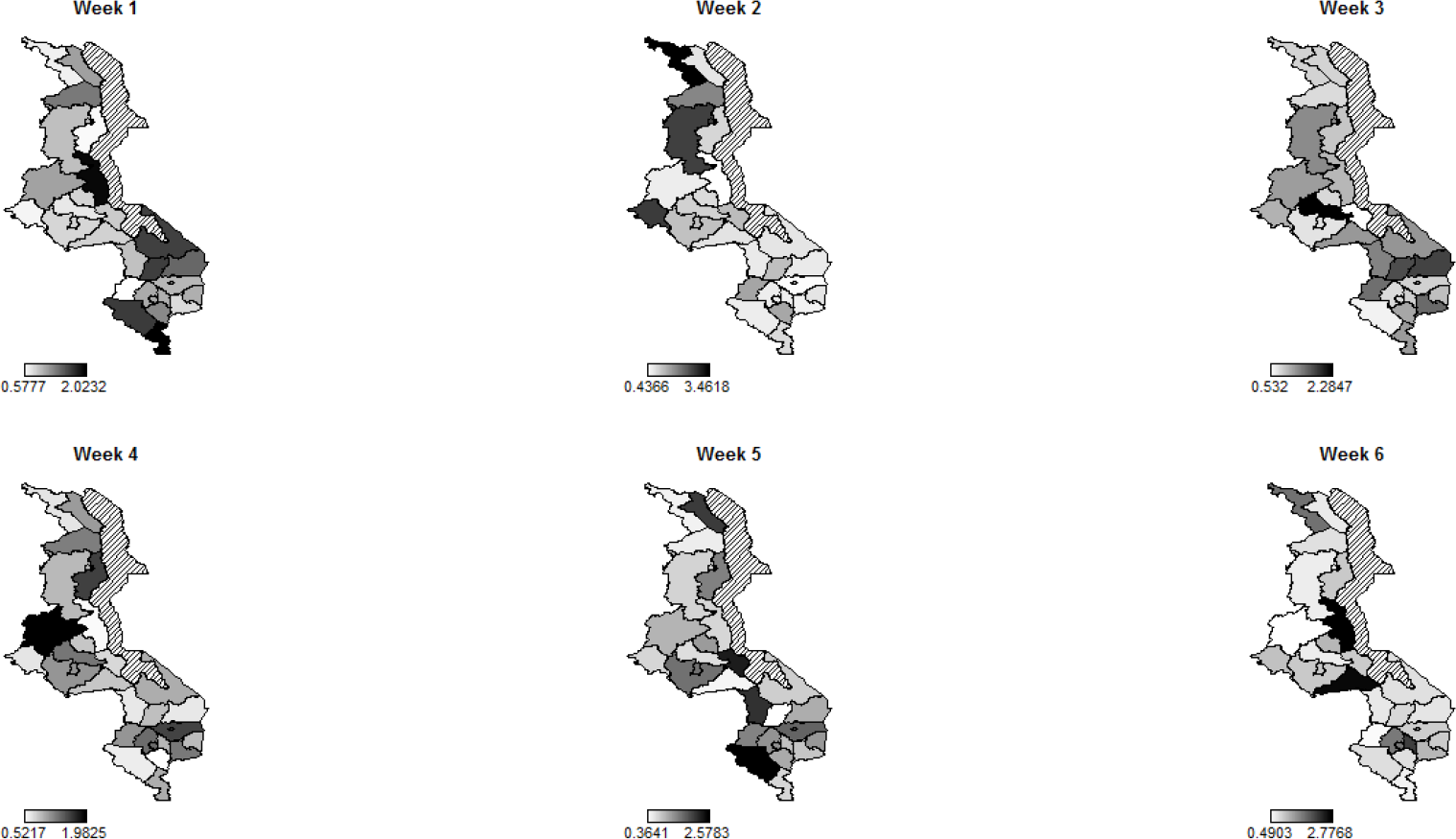
Evolution of spatial risk of COVID-19 over time (Interaction effect of location and time on exponential scale).

## Discussion

The study looked at a snap shot of spatial temporal distribution of COVID-19 in Malawi by focusing on the period, 24^th^ June to 20^th^ August, while using the inseparable statistical spatial temporal model. The use of inseparable model allowed the investigation of the joint or interaction effect of time and location on COVID-19 cases. The use of non-parametric model for time effect (RW2) also enabled the capturing of the subtle influences of time on the risk of contracting COVID-19.

The space distribution of COVID-19 risk in Malawi in the given time period, shows the cities and the surrounding areas being at increased risk. The explanation to the observed spatial gradient is a matter of conjecture. One possible factor driving the observed spatial pattern would be the population size and population density. The cities have higher population density than the rural and COVID-19 is therefore more likely to spread fast through the movement and frequent contact between people. Case comparison investigations have found positive correlation of population density and COVID-19 (Penerliev and Petkov, 2020), where for example in Italy, Lombardy, the population density is three times higher than in Piedmont and the incidence rate was also over three times higher than in Piedmont. Evidence of high population density as a risk of disease transmission has been seen in India where Influenza transmission rates in India have been found to increase above a population density of 282 people per square kilometer (African Centre of Strategic Studies, 2020).

The other possible contributor to the observed rural city spatial gradient of COVID-19 risk in Malawi would be international exposure. In this case, cities have higher international exposure than the rural through international flights among others, which would mean more imported cases. Evidence of international exposure as a risk factor of COVID-19 transmission has been observed in Africa as a whole where countries with high international exposure like South Africa, Nigeria, Morroco, Egypt, and Algeria have had higher COVID-19 cases than their counterparts. International exposure as a fuel of COVID-19 transmission has also been documented in Brazil where it was found that cases increased with increase in international flights jetting into the country (Pequeno et al, 2020). The ability to testing may be another catalyst of COVID-19 risk distribution in Malawi where in the urban centers, a large number of people are tested, hence more are reported to have COVID-19 than in the rural setting. COVID-19 testing rate has been confirmed to be positively correlated with virus infection rate (Hamid et al, 2020).

Regarding the temporal distribution, the disease risk was increasing gently from week 1 to week 4 and increased sharply from week 4 to week 5 when it started to decline in most regions. The relative sharp increase in risk in week 4 and 5 may be attributed to the effects of post presidential general election which was held on 23^rd^ June, 2020. The unrestricted political rallies before the election might have caused a spike in COVID-19 risk thereafter. In addition, the rise in COVID-19 cases during this time would be attributed to the decreasing temperatures at this time of the year as this is the time of cold season. The decline of risk from week 5 to the last week may be due to the increasing temperatures as this time marks the beginning of hot season. Negative correlation between COVID-19 cases and temperature has been documented (Pequeno et al, 2020).

The study did not go without weaknesses. The first weakness was that, due to the absence of population size for each area at each time point, the base population at risk for each area was assumed to constant across time which was not practically valid. The other weakness was that the study did not look at future predictions of COVID-19 risk beyond the specified period of the study to give an idea how the disease would progress thereafter. This would have important implications particularly on planning activities that had been brought to a halt by COVID-19 like education and football games. Nonetheless, the study gave an overview of the disease dynamics in both space and time in the specified time frame so as to identify hot spots in both space and time for further epidemiological investigations or interventions.

## Conclusion

The study found a significant effect of both location and time on COVID-19 risk and the effect of either of the two depended on the other, that is, interaction. The risk of COVID-19 for major cities was high compared to the rural districts and that over time, the risk for rural areas remained relatively lower than in cities. The risk of getting COVID-19 in almost all districts started to decline in the last week which was in August. The implications of the study are that future interventions to halt the disease transmission in case the disease repeats itself, should target the major cities like Blantyre, Zomba, Mangochi, Lilongwe and Mzuzu and that by time, attention should be paid to the month of June and July when it is very cold.

## Data Availability

Data is available upon request from the corresponding author

